## Supplementary Material for "Exploring the gut virome in fecal immunochemical test stool samples reveals novel associations with lifestyle in a large population-based study"

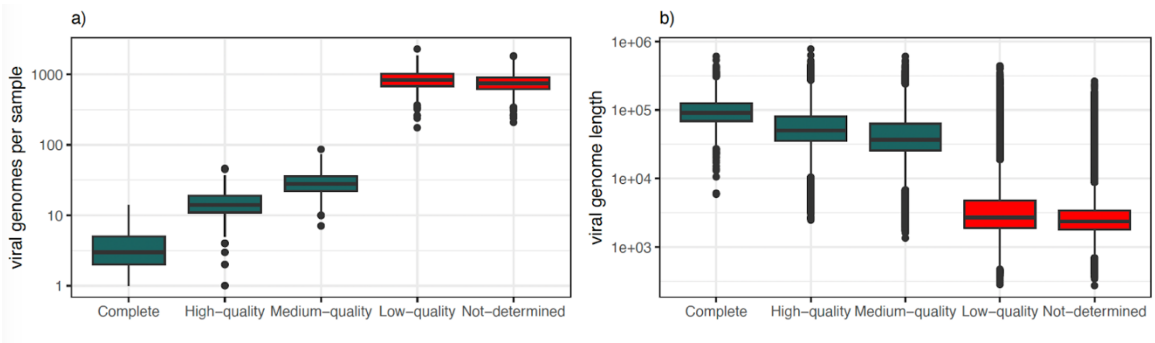

**Supplementary fig. 1 | Quality check of the viral genomes** a) Quality assessment by CheckV revealed that the viral genomes were complete/high-quality, medium-quality and low-quality. b) relationship between the contig length and the genomes quality.

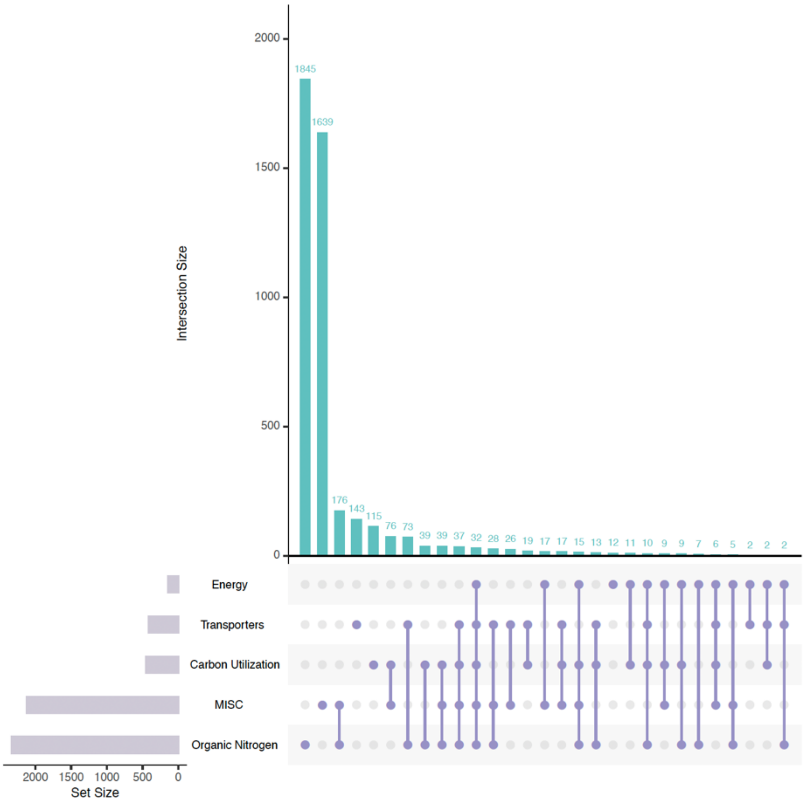

**Supplementary fig.2 | Frequency of AMG detection and intersections in vOTUs according to functional group.** Filled dots with interconnecting vertical lines represent the intersections, and unfilled light gray dots represent sets that do not belong to the intersections. The bars above represent the numbers of AMG's within the intersections, and the bars to the left depict the total number of AMG's in each functional group.

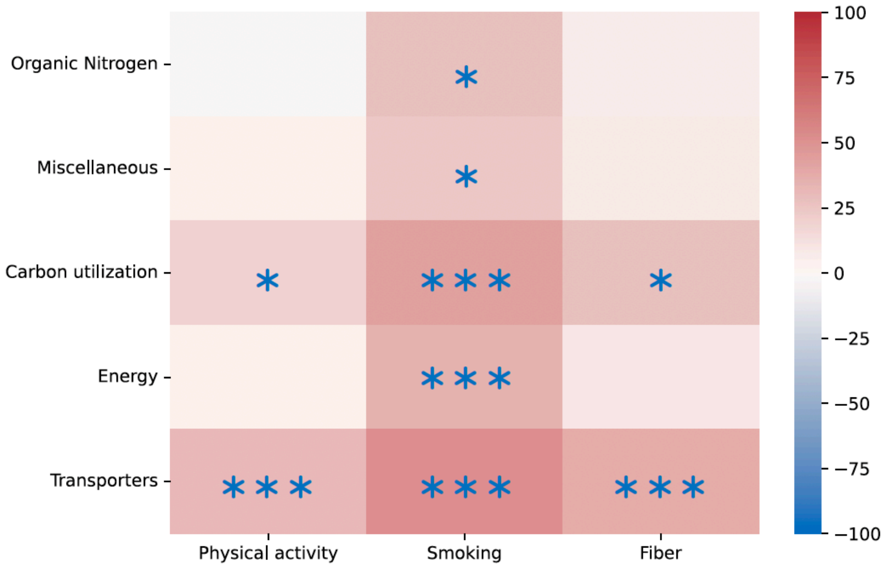

**Supplementary fig.3 | Difference in prevalence of AMGs** in differentially abundant vOTUs compared to their prevalence in all vOTUs. Significance of differences in prevalence were assessed using a binomial test. *p < 0.05, **p < 0.01, ***p < 0.001.

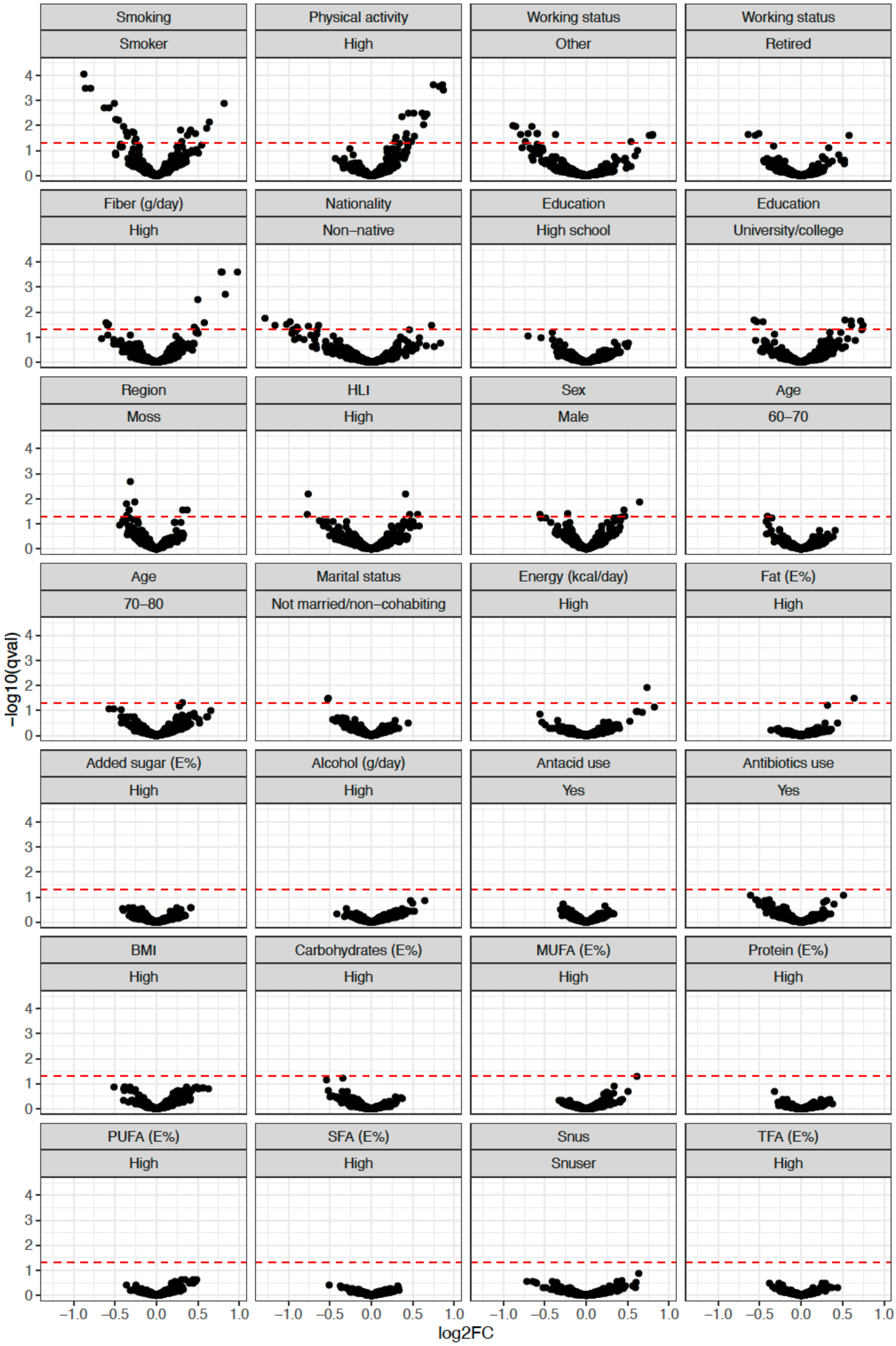

**Supplementary fig.4 | Volcano plots** showing the relationship between effect size (log2 fold change) and significance level (q-value) for vOTUs for diet, lifestyle and demographic variables.

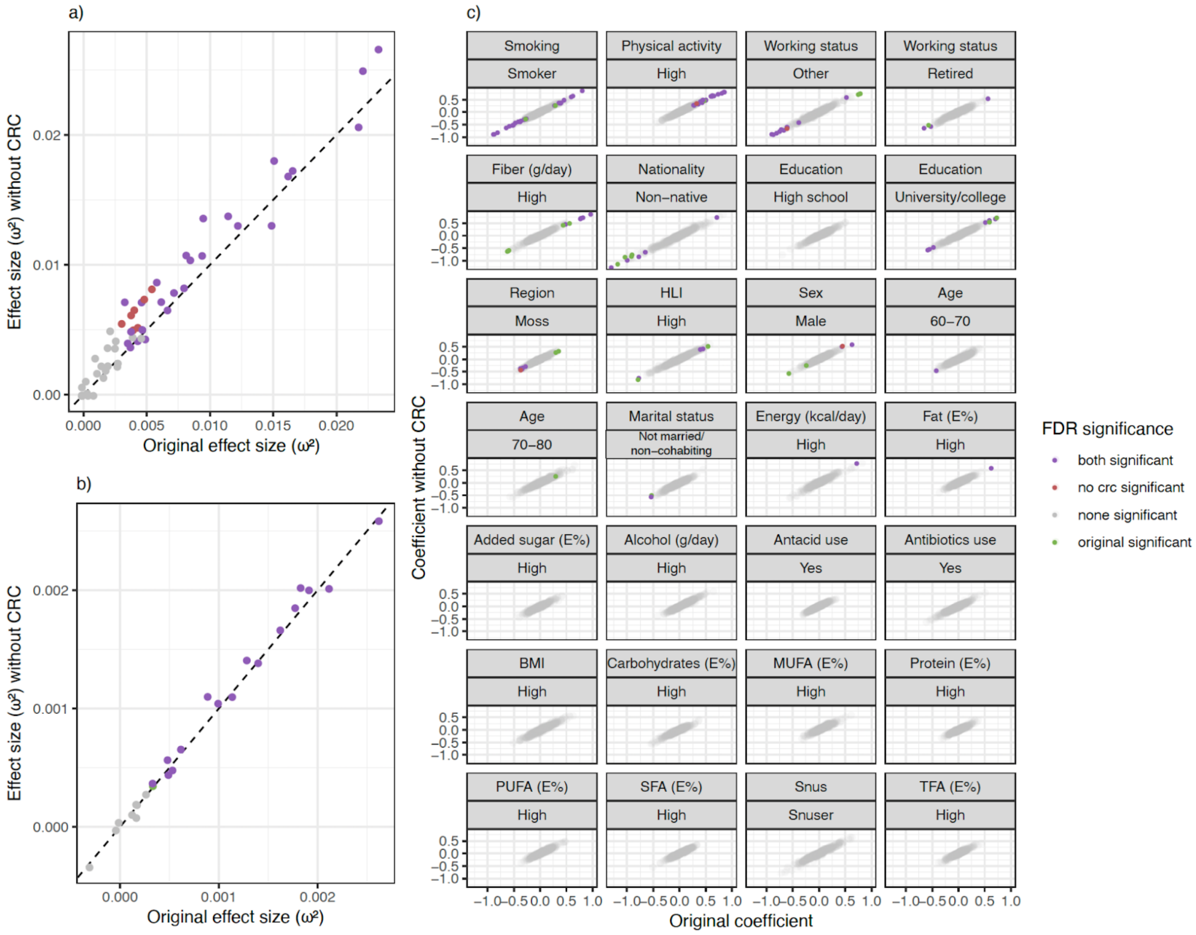

**Supplementary fig. 5 | Sensitivity analyses** excluding any participants with colorectal cancer

**Supplementary Table 1:** Diversity index

| Index | Mean | SD | Median | Min | Max |
|---|---|---|---|---|---|
| Observed | 222.54 | 69.32 | 221 | 41 | 505 |
| Shannon | 4.88 | 0.43 | 4.96 | 2.34 | 7.77 |
| Inverse Simpson | 93.54 | 43.71 | 89.15 | 2.79 | 245 |
| Bray-Curtis dissimilarity | 0.838 | 0.065 | 0.83 | 0.42 | 1 |

**Supplementary Table 2:** Diversity Chao1 Index and fraction by Family

| **Family** | **vOTUs** | **genomes** | **Chao1 index** | **vOTUs/Chao1 index** |
|---|---|---|---|---|
| Microviridae | 528 | 726 | 5340 | 0.0989 |
| Suoliviridae | 161 | 331 | 349 | 0.461 |
| Intestiviridae | 113 | 445 | 196 | 0.577 |
| Steigviridae | 109 | 280 | 229 | 0.476 |
| Peduoviridae | 97 | 132 | 243 | 0.399 |
| Crevaviridae | 33 | 382 | 55 | 0.6 |
| Inoviridae | 32 | 54 | 438 | 0.0731 |
| Winoviridae | 26 | 214 | 35 | 0.743 |

**Supplementary Table 3:** Baseline characteristics of the study participants (n=1034)[1].

| **Demography variables** | |
|---|---|
| Age category, n *(%)* | |
| 50-60 years | 181 (17.5) |
| 60-70 years | 481 (46.5) |
| 70-80 years | 372 (36.0) |
| Male sex, *n (%)* | 582 (56.3) |
| Region, n (%) | |
| Region 1 (Moss) | 568 (54.9) |
| Region 2 (Bærum) | 466 (45.1) |
| Nationality, n (*%)* | |
| Native | 919 (90.5) |
| Non-native | 70 (6.9) |
| Missing | 26 (2.6) |
| Marital status, *n (%)* | |
| Married/cohabiting | 811 (79.9) |
| Not married/non-cohabiting | 202 (19.9) |
| Missing | 2 (0.2) |
| Education, *n (%)* | |
| Primary school | 196 (19.3) |
| High school | 393 (38.7) |
| University/college | 419 (41.3) |
| Missing | 7 (0.7) |
| Working status, *n (%)* | |
| Employed | 322 (31.7) |
| Retired/unemployed | 566 (55.8) |
| Other | 127 (12.5) |
| **Lifestyle variables** | |
| Overall HLI, points | 3.5 (2.8, 4.3) |
| Smoking status, n *(%)* | |
| Non-smoker | 744 (73.3) |
| Smoker | 269 (26.5) |
| Missing | 2 (0.2) |
| Snus status, n *(%*) | |
| Non-snuser | 90 (88.8) |
| Snuser | 70 (6.9) |
| Missing | 44 (4.3) |
| BMI, $kg/m^2$ | 26.5 (24.1, 29.3) |
| Physical activity, min/week | 105 (0, 300) |
| Antibiotic usage, n (*%*) | |
| No | 828 (81.6) |
| Yes | 136 (13.4) |
| Unknown | 51 (5.0) |
| Antacid usage, n (*%*) | |
| No | 701 (69.1) |
| Yes | 269 (26.5) |
| Unknown | 45 (4.4) |

| **Diet variables** | |
|---|---|
| Energy, kcal/day | 2170.4 (1737.3, 2666.1) |
| Protein, E% | 16.6 (15.1, 18.2) |
| Carbohydrates, E% | 41.7 (37.3, 45.8) |
| Added sugar, E% | 4.1 (2.6, 6.5) |
| Fibre, g/day | 27.6 (21.8, 35.1) |
| Fat, E% | 34.5 (31.1, 37.9) |
| SFA, E% | 11.8 (10.2, 13.5) |
| MUFA, E% | 12.8 (11.4, 14.6) |
| PUFA, E% | 6.2 (5.3, 7.3) |
| TFA, E% | 0.3 (0.2, 0.4) |
| Alcohol, g/day | 8.7 (2.4, 18.8) |

*[1]Values are median (p25, p75) for continuous variables and n (%) for categorical variables. The numbers available for analysis vary by variable.*
*Abbreviations: BMI; body mass index, E%; energy percentage, g; gram, MUFA; monounsaturated fatty acids, n; number, PUFA; polyunsaturated fatty acids, SFA; saturated fatty acids, TFA; trans-fatty acids*

**Supplementary Table 4:** Software and Algorithms

| Tool | Version | url |
|---|---|---|
| *Sequence data processing* | | |
| Metagenome-Atlas | 2.4.3 | https://github.com/metagenome-atlas/atlas |
| VirSorter | 2.2.2 | https://github.com/jiarong/VirSorter2 |
| CheckV | 0.8.1 | https://bitbucket.org/berkeleylab/checkv/src/master/ |
| Galah | 0.3.1 | https://github.com/wwood/galah |
| BBMap | 38.96 | https://sourceforge.net/projects/bbmap/ |
| SAM tools | 1.15.1 | https://sourceforge.net/projects/samtools/files/samtools/1.15.1/ |
| vConTACT | 0.11.0 | https://bitbucket.org/MAVERICLab/vcontact2/src/master/ |
| Prodigal<br>DRAMv | 2.6.3 | https://github.com/hyattpd/Prodigal<br>https://github.com/WrightonLabCSU/DRAM |
| graphanalyzer | 1.4.6 | https://github.com/lazzarigioele/graphanalyzer |
| *Workflow management tools* | | |
| Snakemake | | https://snakemake.github.io/ |
| *Visualization tools* | | |
| Cytoscape | v3.7.1 | https://cytoscape.org |
| *Statistics* | | |
| R package vegan | 2.6.2 | https://github.com/vegandevs/vegan |
| R package MaAsLin2 | 1.12 | https://github.com/biobakery/Maaslin2 |
| R package micEco | v0.9.15 | https://github.com/Russel88/MicEco |
| *Databases* | | |
| Pfam | | https://www.ebi.ac.uk/interpro/ |
| CAZy | | http://www.cazy.org/ |
| VOGDB | | https://vogdb.org/ |
| KOfam | | https://www.genome.jp/tools/kofamkoala/ |
| UniRef | | https://www.uniprot.org/ |
| CAN | | https://bcb.unl.edu/dbCAN/ |
| RefSeq | | https://www.ncbi.nlm.nih.gov/refseq/ |
